# Assessing the quality of clinical and administrative data extracted from hospitals: The General Medicine Inpatient Initiative (GEMINI) experience

**DOI:** 10.1101/2020.03.16.20036962

**Authors:** Sachin V. Pasricha, Hae Young Jung, Vladyslav Kushnir, Denise Mak, Radha Koppula, Yishan Guo, Janice L. Kwan, Lauren Lapointe-Shaw, Shail Rawal, Terence Tang, Adina Weinerman, Fahad Razak, Amol A. Verma

## Abstract

**Objective:** Large clinical databases are increasingly being used for research and quality improvement, but there remains uncertainty about how computational and manual approaches can be used together to assess and improve the quality of extracted data. The General Medicine Inpatient Initiative (GEMINI) database extracts and standardizes a broad range of data from clinical and administrative hospital data systems, including information about attending physicians, room transfers, laboratory tests, diagnostic imaging reports, and outcomes such as death in-hospital. We describe computational data quality assessment and manual data validation techniques that were used for GEMINI.

**Methods:** The GEMINI database currently contains 245,559 General Internal Medicine patient admissions at 7 hospital sites in Ontario, Canada from 2010-2017. We performed 7 computational data quality checks followed by manual validation of 23,419 selected data points on a sample of 7,488 patients across participating hospitals. After iteratively re-extracting data as needed based on the computational data quality checks, we manually validated GEMINI data against the data that could be obtained using the hospital’s electronic medical record (i.e. the data clinicians would see when providing care), which we considered the gold standard. We calculated accuracy, sensitivity, specificity, and positive and negative predictive values of GEMINI data.

**Results:** Computational checks identified multiple data quality issues – for example, the inclusion of cancelled radiology tests, a time shift of transfusion data, and mistakenly processing the symbol for sodium, “Na”, as a missing value. Manual data validation revealed that GEMINI data were ultimately highly reliable compared to the gold standard across nearly all data tables. One important data quality issue was identified by manual validation that was not detected by computational checks, which was that the dates and times of blood transfusion data at one site were not reliable. This resulted in low sensitivity (66%) and positive predictive value (75%) for blood transfusion data at that site. Apart from this single issue, GEMINI data were highly reliable across all data tables, with high overall accuracy (ranging from 98-100%), sensitivity (95-100%), specificity (99-100%), positive predictive value (93-100%), and negative predictive value (99-100%) compared to the gold standard.

**Discussion and Conclusion:** Iterative assessment and improvement of data quality based primarily on computational checks permitted highly reliable extraction of multisite clinical and administrative data. Computational checks identified nearly all of the data quality issues in this initiative but one critical quality issue was only identified during manual validation. Combining computational checks and manual validation may be the optimal method for assessing and improving the quality of large multi-site clinical databases.

## BACKGROUND AND SIGNIFICANCE

Routinely collected clinical and administrative health data are increasingly being used in large databases for research and quality improvement.^[1-4]^ In many health systems, the electronic data in hospitals are stored in a complex array of repositories with limited central oversight or standardization. Extracting data from these systems may be prone to errors. Ensuring data quality post-extraction is challenging but important,^[5,6]^ particularly given the impact that large clinical databases might have on quality improvement, research and policy applications.

There are several widely-cited frameworks used to assess data quality.^[7-9]^ A systematic review by Weiskopf and colleagues identified five key data quality dimensions: completeness, correctness, concordance, plausibility, and currency.^[10]^ The methodological approach to examining each dimension remains challenging and poorly described.^[11]^ Manual data validation is often an important step to ensuring data quality.^[12]^ This typically involves manually abstracting a subset of data directly from the source and comparing this to electronically extracted data.^[13]^ However, manual chart reviews are resource-intensive^[13-15]^ and difficult to scale as databases grow in size.^[6]^ Computational data quality assessment has been proposed as an alternative method of ensuring data quality^[16]^ Computational data quality assessment may include application of thresholds to ensure plausibility (e.g. birth date cannot be a future date), visual inspection of linear plots (e.g. to assess temporal trends), and outlier detection formulas.[7,17,18]

Our objective was to describe the combination of computational data quality assessment and manual data validation in a large, real-world, clinical dataset - the General Medicine Inpatient Initiative (GEMINI). This case study offers insights for approaches to ensure data quality in large datasets based on routinely collected clinical data.

## METHODS

### Setting

The General Medicine Inpatient Initiative (GEMINI) database collects administrative and clinical data for all patients admitted to or discharged from the general medicine inpatient service of seven hospital sites affiliated with the University of Toronto in Toronto and Mississauga, Ontario, Canada.^[19]^ The seven hospitals include five academic health centres and two community-based teaching sites. At the time of this manuscript, data had been collected for 245,559 patient visits with discharge dates between April 1, 2010 and October 31, 2017.

GEMINI supports both research and quality improvement applications. The Ontario General Medicine Quality Improvement Network (GeMQIN)^[20]^ uses data from the GEMINI database to create confidential individualized audit and feedback reports for eligible physicians at participating hospital sites.^[21]^ The first version of these reports included six quality indicators (hospital length-of-stay, 30-day readmission, inpatient mortality, use of radiology tests, use of red blood cell transfusions, and use of routine blood-work), and directly informed front-line quality improvement efforts. It was a priority to ensure that high quality data were being used in physician audit-and-feedback reports.

### Ethics

Research Ethics Board approval was obtained from each participating institution.

### Data Extraction, Collection, and Processing

Data extraction occurred over two cycles: 1) patients discharged between April 1, 2010 to March 31, 2015 and 2) patients discharged between April 1, 2015 to October 31, 2017.^[4]^ A wide range of administrative and clinical data were extracted from a variety of data sources (Table 1): patient demographics, diagnostic codes, intervention procedure codes, admission and discharge times, cost of admission, test results from biochemistry, haematology and microbiology laboratories, radiology reports, in-hospital medication orders, vital signs, and blood transfusions. A template document describing requested data formats and standards was provided to each hospital site to facilitate data extraction. Data were extracted by local hospital staff and stored locally in shared network folders.

**Table 1.**
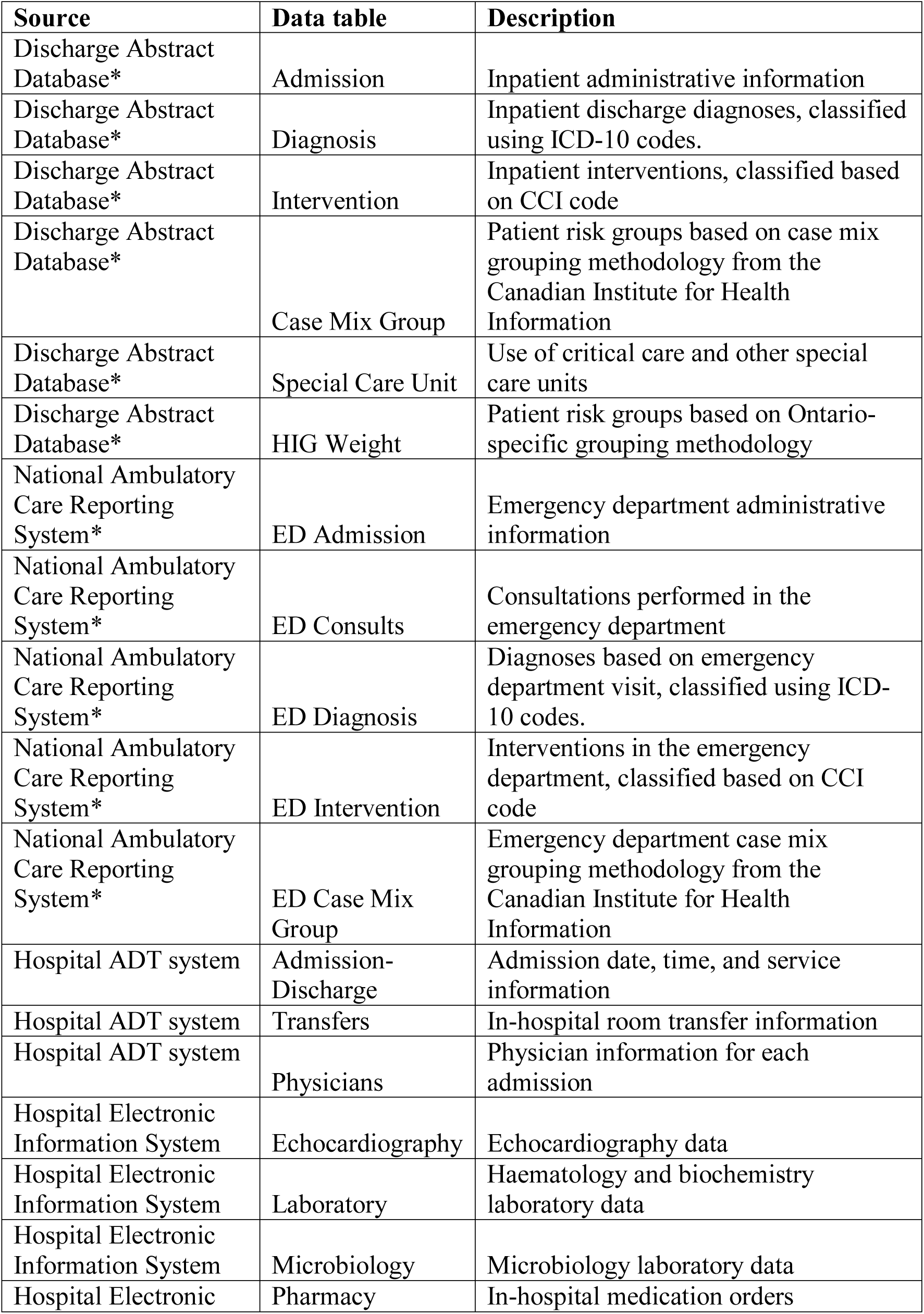
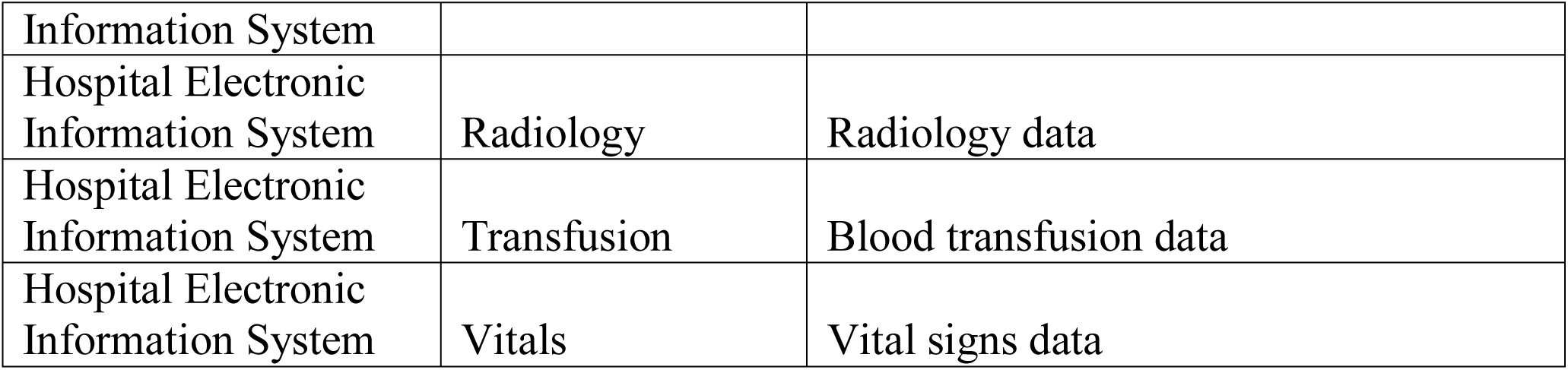
GEMINI database structure. * Data are extracted from hospital sites, based on what each hospital site reports to the Canadian Institute for Health Information for the Discharge Abstract Database and National Ambulatory Care Reporting System. Abbreviations: ADT = admission, discharge, transfer, ICD-10 = 10th revision of the International Statistical Classification of Diseases and Related Health Problems, CCI = Canadian Classification of Health Interventions, HIG = Health Based Allocation Model (HBAM) Inpatient Group, ED = Emergency Department

After data were extracted, GEMINI staff de-identified the data at each hospital site and then securely transferred the data to a central repository. De-identification was performed by removing personally identifying variables from the dataset (e.g. medical record number, encounter number, date of birth, first name, last name, health card number and emergency room registration number).^[22]^ Each patient admission was assigned a unique identifier that allowed for integration of various data tables at the central data repository and re-identification at the local hospital site for the purpose of manual validation.^[23]^ A secure hash algorithm was applied to each patient’s provincial health insurance number, which allowed us to link encounters for the same patient across multiple institutions.^[24]^

In the central repository, GEMINI data were processed and organized into 21 linkable data tables from each hospital site in order to group data into related categories (Table 1). Data were processed to ensure consistent formatting across hospital sites. Data quality was then iteratively assessed by computational data quality assessment and re-extraction as needed, and finally by manual validation.

### Computational Data Quality Assessment

The computational data quality assessments consisted of seven checks. Checks 1-4 were designed to identify errors in data completeness associated with data extraction and transfer procedures and were applied to each data table. Checks 5-7 consisted of detailed inspection of select variables in each table (e.g. sodium tests result values in laboratory data table). Each check assessed different dimensions of data quality (Table 2). The checks are described in detail below:

**Table 2.** Summary of computational data quality checks. Seven computational data quality assessment checks were conducted on entire data tables of specific variables prior to manual validation to assess different dimensions of data quality (reported in a systematic review by Weiskopf and colleagues).^[10]^ We describe each check in detail under the *Methods* section. A data analyst visually reviewed each check to identify issues (unexpected results, outliers, disruptions in trends), which then underwent a step-wise approach including any of data re-extraction, communication with site-specific IT personnel, and manual chart review to correct the issue.

| Check | Name | Level | Description | Quality Dimensions Assessed |
| --- | --- | --- | --- | --- |
| 1 | Patient admissions over time | Data table | Proportion of all GEMINI patient admissions that are present in the data table, by date | Completeness |
| 2 | Data volume over time | Data table | Data volume per data table, by date | Completeness |
| 3 | Admission-specific data volume over time | Data table | Data volume per patient admission, by date | Completeness<br>Plausibility |
| 4 | Distribution of data in relation to admission and discharge times | Data table | Data volume in relation to admission time and discharge time | Completeness<br>Plausibility |
| 5 | Variable presence over time | Variable | Heat map of missing data for each variable, by date | Completeness<br>Plausibility |
| 6 | Specific variable presence over time | Variable | Proportion of data within a data table that pertains to a certain variable after data processing, by date | Completeness<br>Plausibility |
| 7 | Accuracy check | Variable | Categorical variables: report frequencies<br>Numeric variables: report measures of central tendency and distribution | Correctness<br>Concordance<br>Plausibility |

1. **Admissions over time:** We examined the number of patient admissions meeting the inclusion criteria at each hospital site and that were subsequently used to extract data from source systems. We produced a histogram of the proportion of patient admissions that are contained in each data table over time, based on discharge date. This explored whether data for patient admissions was systematically missing from any data table. (Appendix 2 Figure 1)

**Figure 1.**
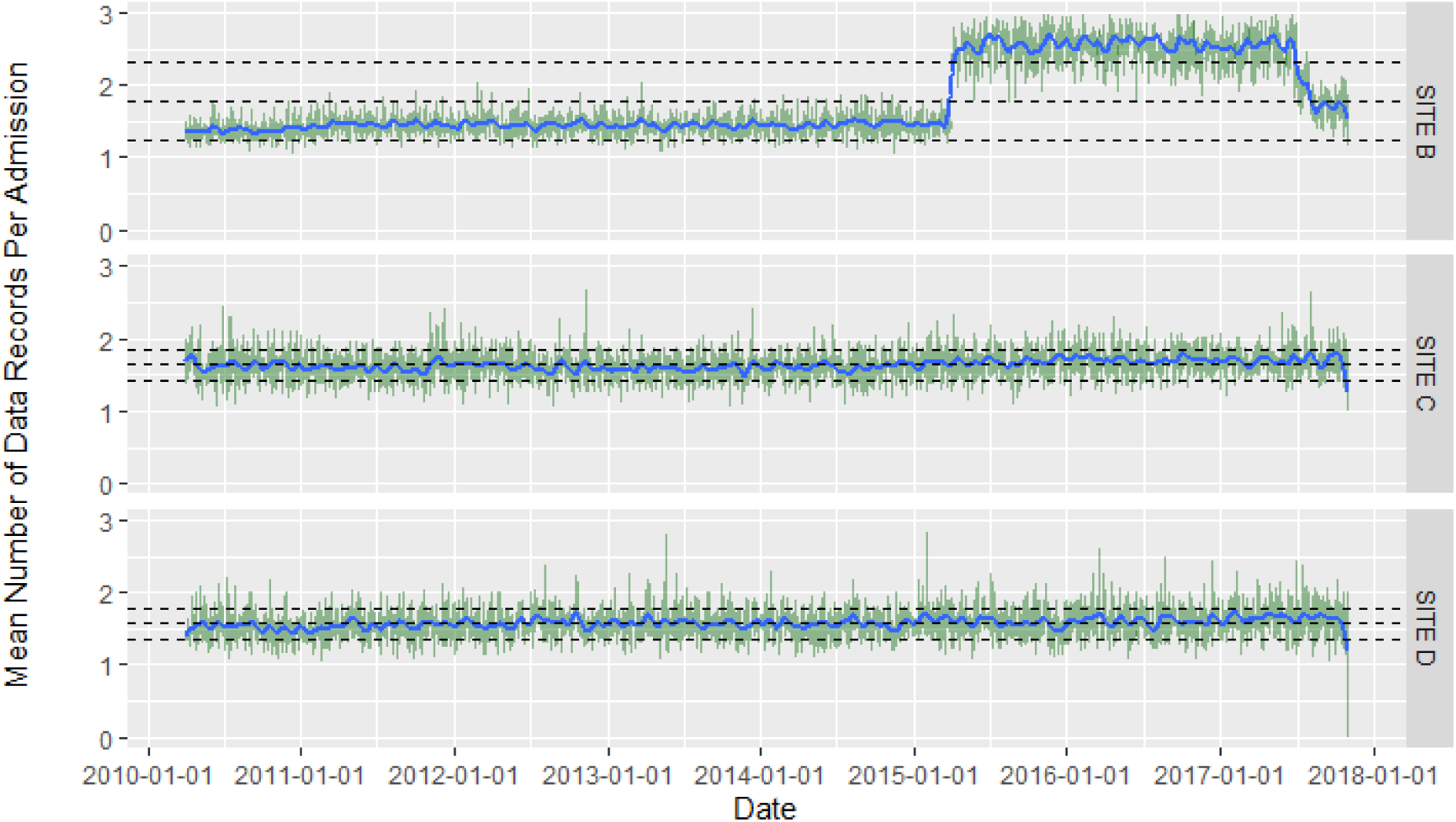
Data volume of radiology tests per patient admission by date at three sites. This is an example of Computational Data Quality Check 3: The x-axis represents the date and the y-axis represents the mean number of radiology tests per patient admission. Sites C and D represent the expected situation, where the number of radiology procedures per patient admission (green lines) and the moving average (blue line) are relatively consistent over time, and within the pre-specified thresholds (bottom and top grey dotted lines represent one standard deviation away from the overall mean). The sharp increase in the average number of tests at Site B between 2015-17 identifies a potential issue, as this level of increase is implausible. Re-extraction did not correct the issue initially. Subsequent manual investigation identified that the extraction had inadvertently included cancelled radiology tests. After successful data re-extraction, without the inclusion of cancelled tests, the aberrancy in the plot was no longer present.
2. **Data volume over time:** We examined the amount of data (as measured by number of rows/observations for each data table) by producing a histogram of total data volume by date and time (if available). In contrast to Check 1, this check assessed missing data, rather than missing patient admissions. Temporal patterns of missingness as well as variations in trends were carefully inspected. (Appendix 2 Figure 2)

**Figure 2A.**
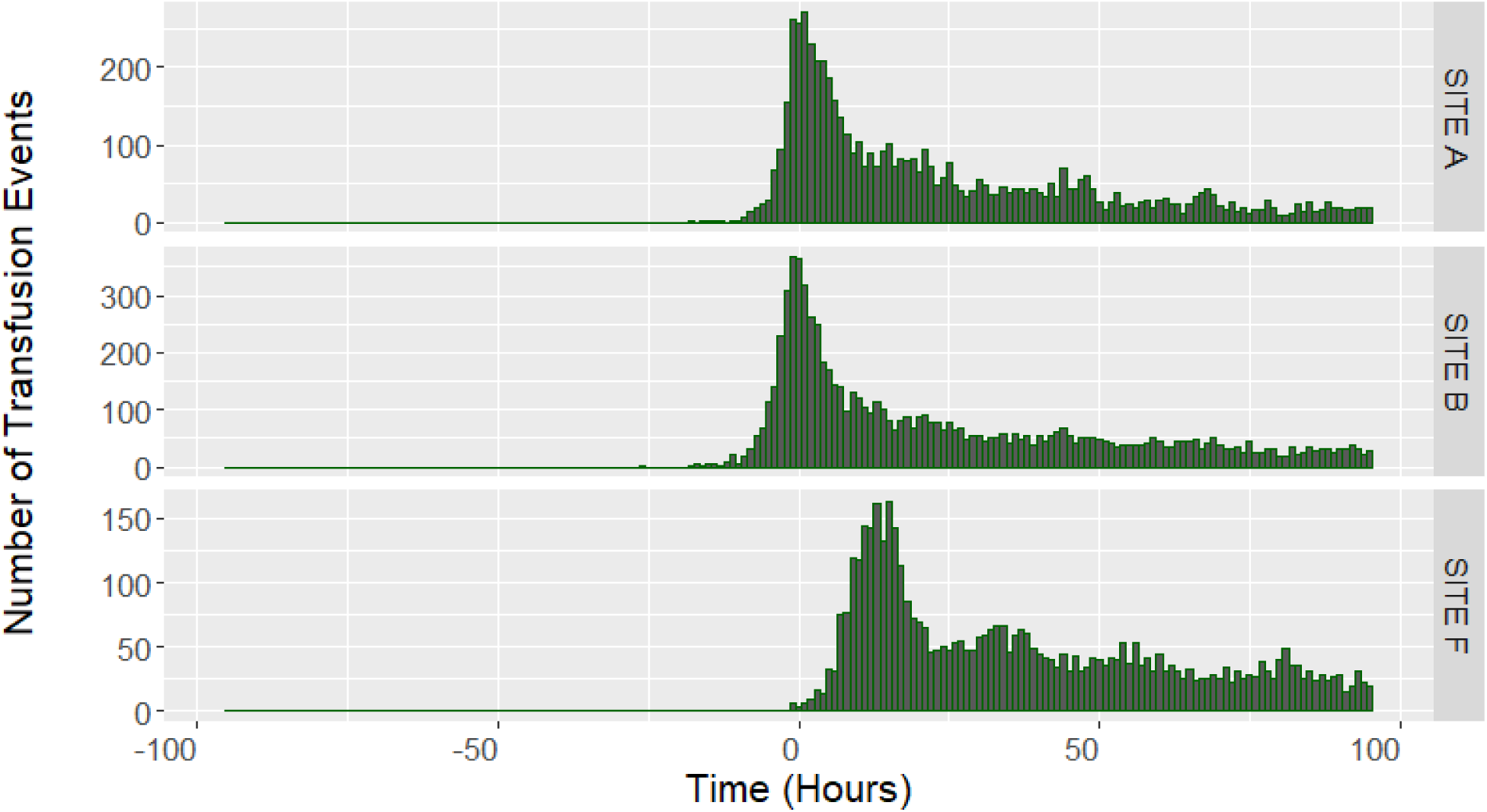
Distribution of the time between transfusion events and the time of admission across three hospital sites.

**Figure 2B.**
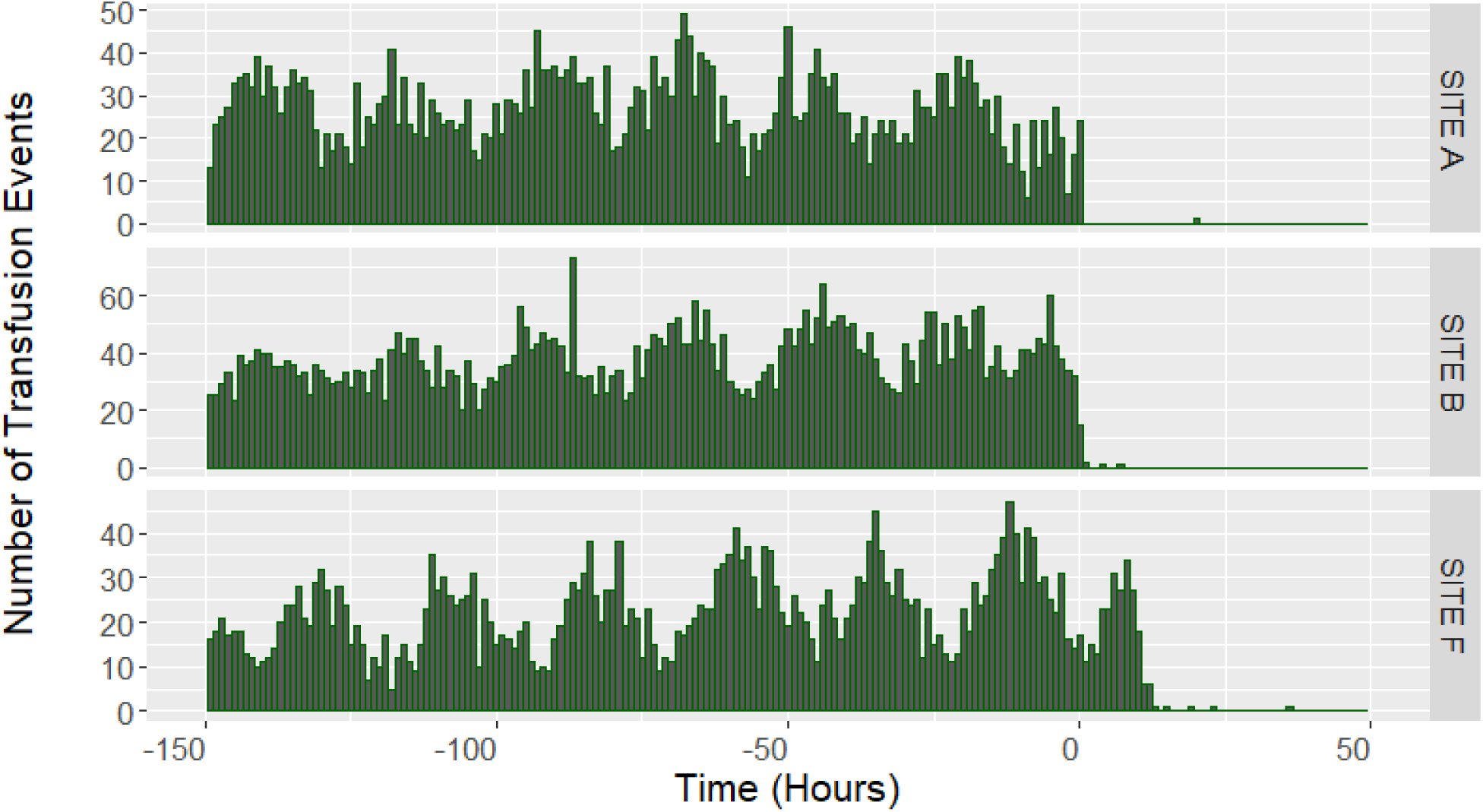
Distribution of the time between transfusion events and the time of discharge across three hospital sites. This is an example of Computational Data Quality Check 4: The x-axis represents the time of the transfusions event in relation to the admission (figure 3A - negative values indicating a time before admission date and time) and discharge (figure 3B – positive values indicate a time after discharge date and time). The y-axis represents the volume of transfusion events that occurred at the time on the x-axis. The plots demonstrate that at Site F, there are no transfusion events prior to admission (at Time<0 Hours). Upon first glance, this suggested missing emergency department transfusion data (i.e. patient in the emergency room and receives a transfusion prior to the admission order) but the issue could not be corrected after re-extraction efforts and discussions with the site data extraction team. The plots also indicate that Site F conducted many transfusions post-discharge (at Time>0 Hours), which would be unlikely. The combination of time from admission, time to discharge, and subsequent manual investigation uncovered a time shift for transfusion data at Site F. This time shift was then corrected in the GEMINI database.
3. **Admission-specific data volume over time:** We examined the total volume of data (as measured by number of rows/observations for each data table) per patient admission to eliminate variation in data volume that may be driven by variations in the number of patient admissions over time. We produced a line graph (with a moving average) of data volume per patient admission by date and time (if available). (Figure 1)
4. **Distribution of data in relation to admission and discharge times:** We examined the distribution of date and time labels on variables in each data table compared to the patient’s admission and discharge date and time (e.g. the difference in time between a radiology test and the patient’s admission date and time). This examined whether date and time information collected were plausible and ensured that we were not missing data from a specific portion of a patient’s hospitalization (e.g. emergency department stay). (Figure 2)
5. **Overall variable presence over time:** We inspected the missingness of every variable from each data table using a heat map. The y-axis was populated with the list of variables in each data table and the x-axis was populated with each row of data in each table, sequenced by admission date. (Appendix 2 Figure 3). This allowed us to identify temporal patterns and clustering in variable missingness.

**Figure 3:**
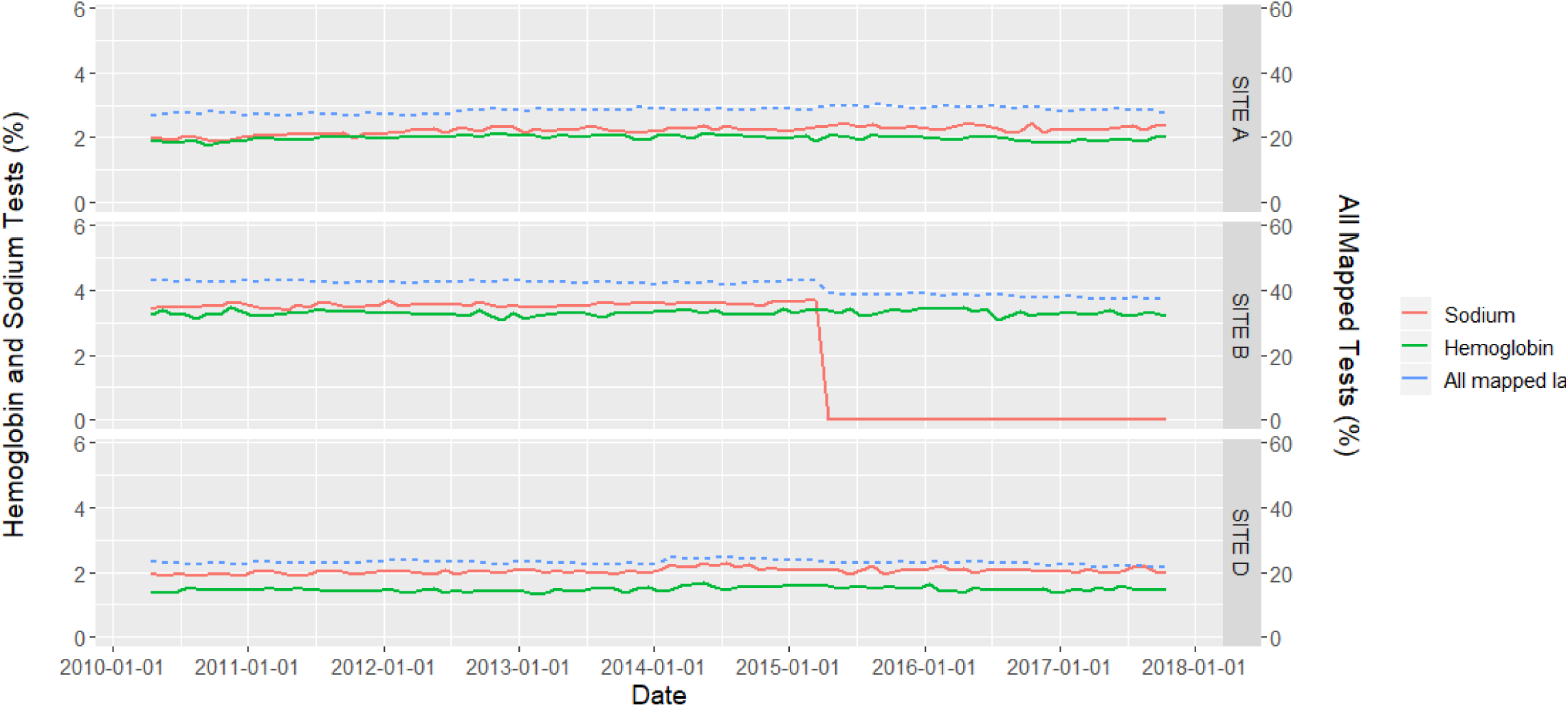
Volume of hemoglobin and sodium laboratory test data over time. This is an example of Computational Data Quality Check 6: The x-axis represents the date. The left y-axis represents the percentage of hemoglobin and sodium tests out of all laboratory tests. The right y-axis represents the percentage of all laboratory data that has been standardized across sites (blue line, many less common laboratory tests had not yet been standardized). There is a sudden decrease in the volume of sodium tests after April 1, 2015, at Site B corresponding to the second round of data extraction. Further investigation revealed that statistical software using default settings mistakenly processed the symbol for sodium “Na” as a null/missing value.
6. **Specific variable presence over time:** We assessed the quality of data categorization and standardization for a specific variable. Because naming conventions differ at each hospital site (e.g. for radiology or laboratory tests) and can change within hospital sites over time, data are manually mapped to categories or standard naming conventions to permit multi-site analysis. For each variable, we produced a line graph of the proportion of mapped data volume at each hospital site by date. Visual inspection of data volume over time allowed us to identify problems with mapping and standardization. (Figure 3)
7. **Plausibility check:** Each variable was inspected to ensure it contained plausible values. For categorical variables, we examined frequency tables. For numerical interval or continuous variables, we computed distributions and measures of central tendency (minimum, mean, median, maximum, and interquartile range). Specific data tables such as laboratory tests required further ad-hoc analysis. For example for each laboratory test type, we inspected the proportion of non-numeric test results, measurement units, and the numerical distribution of test result values.

A data analyst visually reviewed each computational quality check to identify any potential issues of concern. Potential data quality issues could be identified as unexpected results, outliers, and disruptions in trends. These were subsequently investigated through an iterative process. We first examined the original de-identified extracted data table to check if there were any errors with data processing (i.e. formatting, mislabelling, etc.). We next examined the extracted files at each hospital to determine whether there were any errors with deidentification or transfer (i.e. not all files transferred correctly). If the issue remained, we then worked with the hospital site to re-extract the problematic data and we then performed the computational data quality check again. If the issue persisted, a data abstractor manually reviewed data of individual problem cases directly from the patient records in the hospital electronic information system to identify differences between extracted data and the electronic information system. They then reported these differences to the local hospital site IT personnel to help fix the data quality issue. These processes typically led to correction of the data quality issue, after which time the computational data quality check was repeated. If these steps did not correct the data quality issue, the data remained in the GEMINI database but that specific data table was flagged such that it would not be used to inform any quality improvement efforts or academic research. Computational data quality checks occurred in the central data repository and did not require direct access to the source data at individual hospital sites.

### Manual Data Validation

Manual data validation was performed on the GEMINI database that had been improved based on computational data quality assessment and iterative re-extraction from hospitals. GEMINI data were compared to data that could be accessed through each hospital’s electronic medical record (i.e. the information that clinicians see when providing care), which was taken to be the gold standard. We focused primarily on data that were included in the physician audit-and-feedback reports. Thus, we manually validated data from six data tables (laboratory, radiology, physicians, admission, transfers, and transfusion). Two cycles of manual validation occurred, corresponding to each cycle of data extraction. Because two hospital sites share a single electronic information system, we grouped these two together and therefore refer to six hospital sites (A-F) for manual data verification.

A data abstractor manually reviewed data for a sample of patient admissions at each of the six sites. The data abstractor was not given access to the patient admission data stored in the GEMINI database meaning that they were “blinded” to any expected results. For 2010-2015 data, admissions were sampled randomly and with a more targeted approach to ensure sufficient sampling of rare events (Appendix 1):

- 100 random admissions per site for laboratory data table (specific variables were haemoglobin, sodium, creatinine, calcium, aspartate transaminase, international normalized ratio, and troponin)
- 100 random admissions per site for radiology data table (specific variables were computed tomography, plain radiography, ultrasound, magnetic resonance imaging, and echocardiography)
- 200 random admissions per site for physician data table (specific variables were admitting physician name and discharging physician name)
- 800 selected admissions per site for death, transfers, and transfusions data tables (specific variables being mortality, critical care unit transfer, and red blood cell transfusions, respectively).

Because we performed extensive manual data validation in the first cycle, we adopted a more targeted approach in the second cycle (2015-2017 data). For the second cycle, data were manually abstracted from a random sample of 5 admissions for each physician who would be receiving an audit-and-feedback report (approximately 20-30 physicians at each site corresponding to 100-150 admissions at each site).

The data abstractor recorded the result and date and time for the first (or only) occurrence of each specific variable to be validated in the hospital’s electronic medical record for each patient admission. This was considered the gold standard and was compared against the data in the GEMINI database. To avoid potential human recording errors, the data abstractor double-checked the electronic medical record in the event of a discrepancy between the GEMINI database and the manually recorded data. We report both cycles of data validation together. We calculated the sensitivity, specificity, positive predictive value, and negative predictive value of GEMINI database for each data table and for individual variables, overall and stratified by hospital site. We also calculated the overall accuracy (true values / total values) for each data table. Analyses were performed using R version 3.5.2.

## RESULTS

### Computational Data Quality Assessment

Each of the seven computational data quality assessment checks identified data quality issues, many of which were subsequently corrected (Table 3). For example, check 3 (admission-specific data volume over time) identified a sharp implausible increase in radiology tests at Site B that resulted from the inappropriate inclusion of cancelled radiology tests in extracted data (Figure 1). Check 4 (distribution of data with admission and discharge times) identified an implausible time shift of transfusion data at Site F, which made it appear as if patients received transfusions after discharge (Figure 2). Check 5 (variable presence over time) identified a data processing and mapping issue whereby the raw hospital sodium test code was interpreted as null or missing during the second cycle of data extraction. This occurred because the abbreviation, “NA” (Figure 3), which refers to sodium in chemical notation, was interpreted as missing by our statistical software.

**Table 3.** Examples of errors identified using computational data quality checks. Each computational data quality assessment check identified a data quality issue, the causes of which were determined to be data extraction/transfer, data processing, or hypothetically local de-identification (though we did not find an example of this). Some issues that were identified by the computational checks required a manual chart review to fix, while others did not.

| **Quality Check** | **Example of Data Quality Issue Identified** | **Cause of Issue** | **Manual chart review required to investigate and fix** |
| --- | --- | --- | --- |
| 1. Patient admissions over time | Missing ED data extraction for certain patient admissions in 2015 at one site (Appendix 2 Figure 1) | Data extraction/transfer | No |
| 2. Data volume over time | Missing transfusion data during two time periods at one institution (Appendix 2 Figure 2) | Data extraction/transfer | No |
| 3. Admission-specific data volume over time | Incorrectly included cancelled radiology tests resulting in sharp increase in radiology tests at one institution (Figure 1) | Data extraction/transfer | Yes |
| 4. Distribution of data in relation to admission and discharge times | Transfusion data at one site was time-shifted, leading to the timestamp on certain transfusions being after patient discharge (Figure 2) | Data extraction/transfer | Yes |
| 5. Overall variable presence over time | Glasgow Coma Scale data at one institution was missing (Appendix 2 Figure 3) | Unclear cause at this time | Yes |
| 6. Specific variable presence over time | At site B, “NA” was used to describe sodium tests. At the data processing stage, statistical software recognized this as a null/missing value. (Figure 3) | Data processing at central repository | No |
| 7. Plausibility check | No quality issues identified | Data extraction/transfer | No |

When data quality errors were identified by computational checks, we attempted to determine which step of the data extraction, collection, and standardization process was the source of the error. Computational data quality checks identified errors arising from data extraction/transfer and processing (Table 3) and could hypothetically identify errors that occur during local de-identification, though we did not find an example of this.

### Manual Data Validation

Across all sites, data tables, and both cycles of manual validation, 23,419 data points were manually abstracted from 7,488 patient admissions. The specific number of data points for each data table and each specific variable at each site is listed in Appendix 2.

Compared to the gold standard of data manually abstracted from an electronic medical record, the GEMINI database was found to be highly sensitive (Table 4), ranging from 95%-100% across data tables, and highly specific, ranging from 99%-100% across data tables. The database was also found to have high positive and negative predictive values, with overall results ranging from 93-100% and 99-100%, respectively, across data tables. The overall accuracy of the database was found to be 98%-100% across data tables.

**Table 4.**
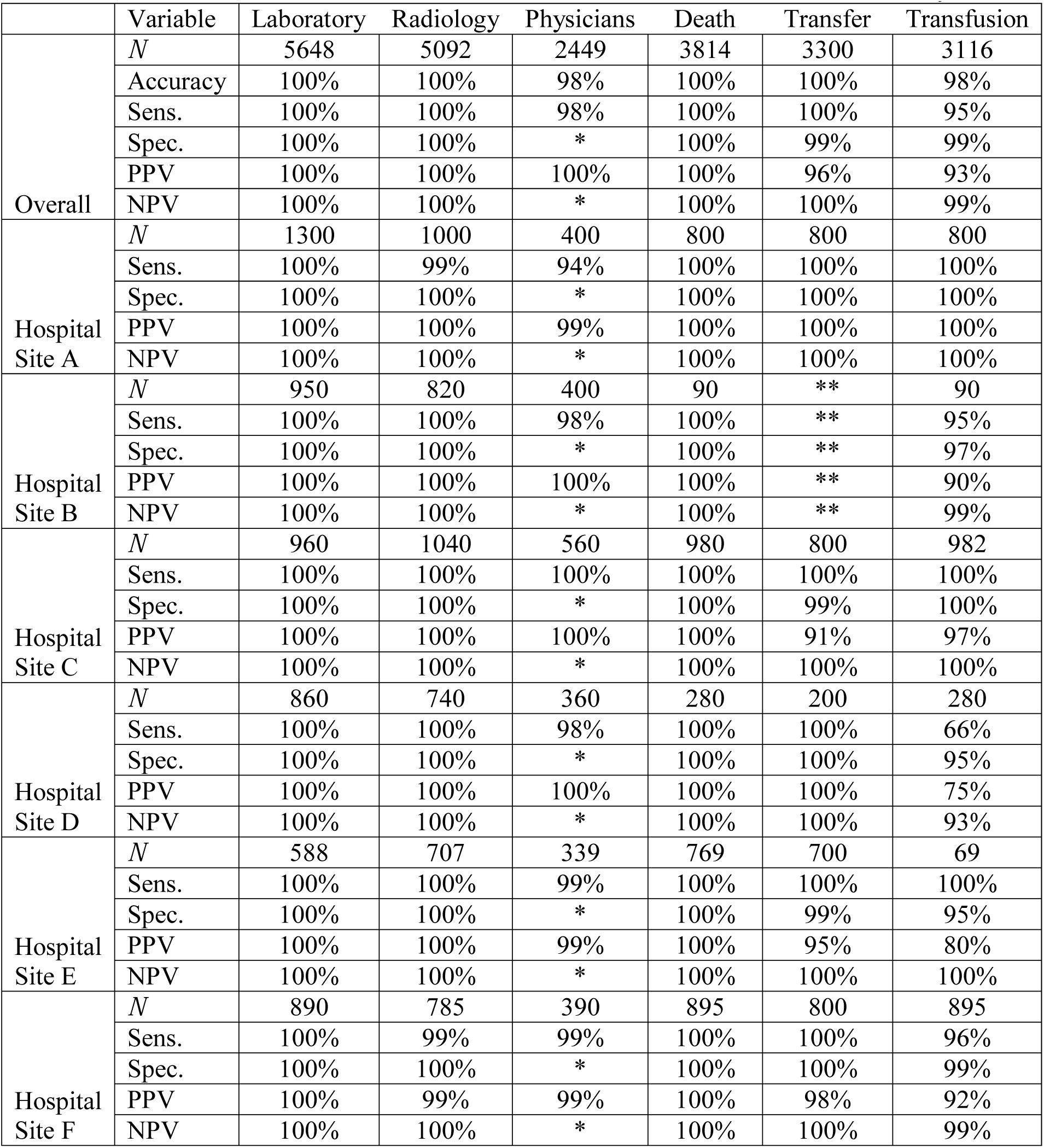
Manual data validation results for each data table, overall and stratified by site. N=Number of data points, which is distinct from number of patient admissions. For example, at site A, 1000 radiology data points were manually checked, but these came from 200 patient admissions for each type of radiology test. *For physician variables (accuracy of admitting and discharging physician names), specificity and negative predictive value cannot be calculated given the lack of true negatives (i.e. every individual should theoretically have an admitting and discharging physician, making the number of true negatives equal to 0). **Site B used paper charts for critical care transfer thus manual data validation was not feasible.

Manual data validation identified one important data quality issue that was not flagged by computational approaches. Specifically, blood transfusion data at Site D had poor sensitivity (66%) and positive predictive value (75%), because of problems with the date and time that the transfusions reportedly occurred.

## DISCUSSION

This paper reports the experience of an extensive data quality assessment effort involving a broad range of administrative and clinical data extracted from 7 hospital sites over 8 years. We highlight a feasible and pragmatic approach to computational data quality assessment and illustrate how various data quality issues were identified. After manual validation of over 23,000 data points, the GEMINI database was found to have an overall accuracy of 98%-100% compared with hospital site records. Our experience suggests that although computational data quality checks are effective, they may not identify all important data quality issues. Specifically, we identified crucial data quality issues in blood transfusion data at one hospital site that were not detected through computational data quality checks. The GEMINI experience suggests that computational and manual approaches should be used together to iteratively improve and validate databases that are extracted from clinical information systems.

Data quality assessment is crucial before routinely collected data can be used for secondary purposes, such as research or quality improvement.^[25]^ One flexible approach is to ensure that data are “fit-for-purpose” by data consumers, which has informed numerous frameworks and models for data quality assessment.^[7,9,10,26,27]^ However, less has been published about how to operationalize these approaches, particularly in multi-site clinical datasets.^[7,28,29]^ Kahn and colleagues describe a conceptual model and a number of computational rules to explore data quality in electronic health record-based research.^[7]^ Similarly, van Hoeven and colleagues articulate an approach to assessing the validity of linked data using computational methods and report its application in a specific case using transfusion data.^[28]^ Terry and colleagues developed 11 measures of quality for primary health care data extracted from electronic medical records.^[29]^ Each of these studies admirably documents the process of operationalizing conceptual data quality frameworks into real-world applications. These studies all focus on computational quality checks. Conversely, Baca and colleagues report a data validation effort of the Axon Registry, a clinical quality data registry, and focus entirely on the validation effort but do not describe computational efforts to assess and improve data quality within the registry.^[30]^ We have been unable to find any studies that report the experience of using both computational and manual approaches to data quality assessment. Our study extends the literature by operationalizing broad data quality domains, specifically using computational data checks to inform iterative data quality improvement, and reporting the effectiveness of this approach based on rigorous manual validation across a range of data types and healthcare organizations.

The main implication of our study is that computational quality checks can identify most data quality issues but not all. Blood transfusion data in our study highlight the strengths and weaknesses of computational checks. Computational checks identified that transfusion data were time-shifted at one hospital, because some blood transfusions were apparently administered after patient discharge, which was implausible. However, computational checks missed major inaccuracies in the dates and times of blood transfusions at another hospital because the errors were not systematic and did not create any discernible patterns. The GEMINI experience suggests that although manual data validation is labour-intensive, it may be necessary to ensure high quality data. Starting with computational checks and targeting manual data validation with a “fit-for-purpose” strategy (as we did in our second data collection cycle when we validated a smaller sample based on physician quality reports) may minimize manual workload without sacrificing data quality.

Our study is limited in several ways. First, we only performed manual validation on certain data tables, stemming from our “fit-for-purpose” approach focused on audit-and-feedback reports.^[26]^ Given that this included a breadth of data tables, we feel that our findings are likely generalizable to other data tables within GEMINI. Second, although we describe the need to manually review the visual computational data quality checks, we were unable to quantify the labour hours required for either computational quality assessment or manual validation. Third, our approach to manual data validation cannot address data quality issues at the source system (e.g. missing values or incorrect data entry) as these are already embedded into the data, which form our ‘gold standard’. Finally, we focused on traditional domains of data quality, but future research could further assess non-traditional aspects such as context, representation, and accessibility.^[27]^

## CONCLUSION

The GEMINI experience highlights the importance of an iterative data quality assessment methodology that combines computational and manual techniques. Through this approach, GEMINI has achieved highly reliable data extraction and collection from hospital sites. Our findings demonstrate that computational data quality assessment and manual validation are complementary and combining these should be the ideal method to assess the quality of large clinical databases. Future research should focus on methods to reduce the amount of manual validation that is needed, and to assess non-traditional aspects of data quality.

## Data Availability

The data used to create this manuscript is available in the GEMINI database.

